# A Deep Attention-Based Encoder for the Prediction of Type 2 Diabetes Longitudinal Outcomes from Routinely Collected Health Care Data

**DOI:** 10.1101/2024.11.02.24316561

**Authors:** Enrico Manzini, Bogdan Vlacho, Josep Franch-Nadal, Joan Escudero, Ana Génova, Elisenda Reixach, Erich Andrés, Israel Pizarro, Dídac Mauricio, Alexandre Perera-Lluna

## Abstract

Recent evidence indicates that Type 2 Diabetes Mellitus (T2DM) is a complex and highly heterogeneous disease involving various pathophysiological and genetic pathways, which presents clinicians with challenges in disease management. While deep learning models have made significant progress in helping practitioners manage T2DM treatments, several important limitations persist. In this paper we propose DARE, a model based on the transformer encoder, designed for analyzing longitudinal heterogeneous diabetes data. The model can be easily fine-tuned for various clinical prediction tasks, enabling a computational approach to assist clinicians in the management of the disease. We trained DARE using data from over 200,000 diabetic subjects from the primary healthcare SIDIAP database, which includes diagnosis and drug codes, along with various clinical and analytical measurements. After an unsupervised pre-training phase, we fine-tuned the model for predicting three specific clinical outcomes: i) occurrence of comorbidity, ii) achievement of target glycaemic control (defined as glycated hemoglobin *<* 7%) and iii) changes in glucose-lowering treatment. In cross-validation, the embedding vectors generated by DARE outperformed those from baseline models (comorbidities prediction task *AUC* = 0.88, treatment prediction task *AUC* = 0.91, HbA1c target prediction task *AUC* = 0.82). Our findings suggest that attention-based encoders improve results with respect to different deep learning and classical baseline models when used to predict different clinical relevant outcomes from T2DM longitudinal data.

## 1. Introduction

It is estimated that 529 million people worldwide have Type 2 Diabetes Mellitus (T2DM), and current evidence suggests that this prevalence will exceed 1.3 billion by 2050[1]. Despite being a highly heterogeneous disease with variable progression patterns and risks of comorbidities, T2DM is primarily diagnosed and treated based on a single metabolite, namely glucose[2]. The usage of Machine Learning (ML) could reshape the paradigm of clinical care, enhance efficiency, and lead to a greater emphasis on patient-centered approaches[3]. In particular, Deep Learning (DL) techniques have been shown to get better results compared to traditional approaches for different tasks and using different types of clinical data: from disease detection to sequential prediction of clinical events; from data augmentation to concept embedding[4]. In the field of diabetes, DL algorithms exhibit a diverse range of applications encompassing the diagnosis and prediction of diabetes onset, glucose management, and the forecasting of diabetes-related comorbidities[5, 6], such as diabetic retinopathy[7] or cardiovascular complications[8].

In this study we proposed DARE (Diabetic Attention with Relative position Representation Encoder), an attention-based encoder for the analysis of T2DM evolution. DARE is a deep learning framework that can be used to manage various sub-goals related to the management of the T2DM. The contributions of this model are manifold: (1) we analyse sequences of multi-modal clinical events, including diagnosis and drugs codes, but also different clinical and analytical continuous variables without the need to categorize them; (2) we introduce in the input sequence a state vector describing information about the subject that helps the model to better learn the sequence of events; (3) we incorporate a Relative Position Representation (RPR) attention layer [9], in order to better learn the irregularity of the data. We validated the model on three different diabetes-related clinical tasks, namely: occurrence of diabetes-related comorbidities; changes in glucose-lowering treatments; and glycated hemoglobin (HbA1c) target prediction. DARE’s performance surpassed other deep learning and classical baseline models for all three outcomes.

## 2. Related Work

A major challenge in training deep learning (DL) models for specific tasks is the limited availability of labeled data for training and validation. For this reason, the concept of transfer learning, i.e. training a model on a generic domain to transfer this knowledge on a different, more specific, one,[10] is gaining increased traction in DL research. Among architectures for transfer learning, the models based on the Transformer architecture[11] are particularly effective, using self-attention mechanisms to capture long-range dependencies and contextual relationships in sequential data. Specifically, the Bidirectional Encoder Representations from Transformers (BERT) model, a Transformer encoder architecture, has achieved state-of-the-art results across a range of Natural Language Processing (NLP) tasks by effectively modeling complex, lengthy sequences[12].

This capability has led to significant interest in adapting the Transformer encoder to other domains outside NLP, including the clinical domain. For instance, few studies have applied Transformer-based methods to electronic health record (EHR) data to predict disease occurrences. Med-BERT, for example, was proposed to predict pancreatic cancer and heart failure in diabetes patients, leveraging data from 600 U.S. hospitals to achieve AUC improvements of 1.62–6.14% over baseline models[13]. Similarly, BEHRT was pre-trained on a large EHR dataset, demonstrating superior predictive power for conditions like epilepsy, prostate malignancy, and depression, while its extension, Hi-BEHRT, achieved even better performance for predicting heart failure, diabetes, chronic kidney disease, and stroke[14, 15].

Despite the progress, to the best of our knowledge, there are no appications of Transformer-based models specifically predicting complications in individuals with type 2 diabetes mellitus (T2DM), while the application of other DL methods are limited: Dworzynski et al., for example, used logistic ridge regression, random forest, and decision tree-gradient boosting on Danish National Patient Register data outperforming logistic regression models with AUCs from 0.69 to 0.80 in the prediction of heart failure, myocardial infarction, stroke, other cardiovascular diseases, and chronic kidney disease[16]. Similarly, Ravaut et al. employed decision tree-gradient boosting to predict various adverse outcomes in T2DM patients, with an average test AUC of 0.77[17].

Also in the context of glucose-lowering treatment prediction, no studies have yet utilized Transformer encoders. However, Shang et al.’s G-BERT model, applied to a related task of medication recommendation, achieved promising results (AUC 0.66–0.69), highlighting the potential of Transformer-based models for treatment prediction in T2DM[18]. Our study aims to build on this potential, specifically exploring the use of Transformer encoders for predicting glucose-lowering medication outcomes in T2DM.

For glycemic control prediction, Nagaraj et al.’s 2019 study developed a supervised machine learning model to predict HbA1c response after insulin treatment in 1,188 T2DM patients, achieving AUC values of 0.80 or greater. However, the study faced limitations related to patient characteristics and control over confounding variables[19].

## 3. Material and Methods

### 3.1. Study design

Data from this study were extracted from the Information System for the Development of Research in Primary Care (SIDIAP) database[20]. The SIDIAP database contains data from electronic health records (EHRs) collected from approximately 5.6 million patients registered from 287 Primary Care Centres (PCC) in Catalonia (Spain). It comprises data on patient demographics, health problems, visits to healthcare professionals, clinical variables, prescriptions and dispensations of medication, and laboratory test results from routine health surveillance and health care. This data covered five full calendar years, from 2013 to 2017.

This analysis included only subjects with a confirmed diagnosis of T2DM, defined by having at least one International Classification of Diseases, Tenth Revision (ICD-10) code from groups E11 or E14 [21]. Additionally, we excluded subjects under 18 years old and those with any codes for type 1, secondary, or gestational diabetes. Further details on data extraction and subject selection criteria were published previously [22].

Upon inclusion in the study during the first year (2013), the following clinical and laboratory variables related to diabetes were available: glycated hemoglobin (HbA1c), body mass index (BMI), diastolic and systolic blood pressure (DBP and SBP), high-density lipoprotein (HDL), low-density lipoprotein (LDL), total cholesterol, triglycerides (TG), creatinine, albumin to creatinine ratio (ACR), Glomerular filtration rate (eGFR) estimated with CKD-EPI and MDRD methods, the Framingham-REGICOR estimation score for the coronary risk [23], and the ankle-brachial Index (ABI). Additionally, we collected data on Glucose-lowering medications: prescriptions belonging to the A10 group of the Anatomical Therapeutic Chemical Classification System (ATC) [24], and Diagnoses of the most common diabetes comorbidities: hypertension (HTN), cardiovascular disease (CVD), neuropathy, retinopathy, and chronic kidney disease (CKD). Specific ICD-10 codes used for these diagnoses are provided in Supplementary Material, Table S1.

### 3.2. Data Representation and Model Development

DARE is an encoder model based on the Transformer encoder architecture, designed to analyze sequences of clinical records beginning from an initial time point. For each subject, denoted as *p*_*j*_, the model takes as input their initial status *P*_*j*_(*t*_0_) (divided into three vectors *P*_*j,gen*_, *P*_*j,D*_(*t*_*i*_), *P*_*j,T*_(*t*_*i*_) representing general static information of the patient, diagnosis, and treatments) and a sequence of routinely collected healthcare data (RCHCD) 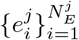, which includes electronic health record (EHR) data gathered as part of routine clinical operations. Each event in this sequence is represented as a triplet, specifying the type (e.g., diagnosis, measured variable, new prescription), the measured value (if applicable), and the time of the event.

DARE architecture is shown in Figure 1. The model operates with two subsequent layers: the first layer is an embedding layer that transforms the original input data (event type, value, and time) into fixed-size vectors. This embedding approach is similar to the one used in the HE-LSTM model[25]. Here, each vector is the sum of the embeddings for the event type, value, and time; the second layer is a modified transformer encoder that relates these vectors to each other through attention mechanisms. To account for the varying time intervals between events in the sequence, we incorporate a relative position representation into the self-attention mechanism, following the approach of Shaw et al.[9]. This allows the model to consider the distance between events. Further details on model implementation can be found in Supplementary Material.

**Figure 1:**
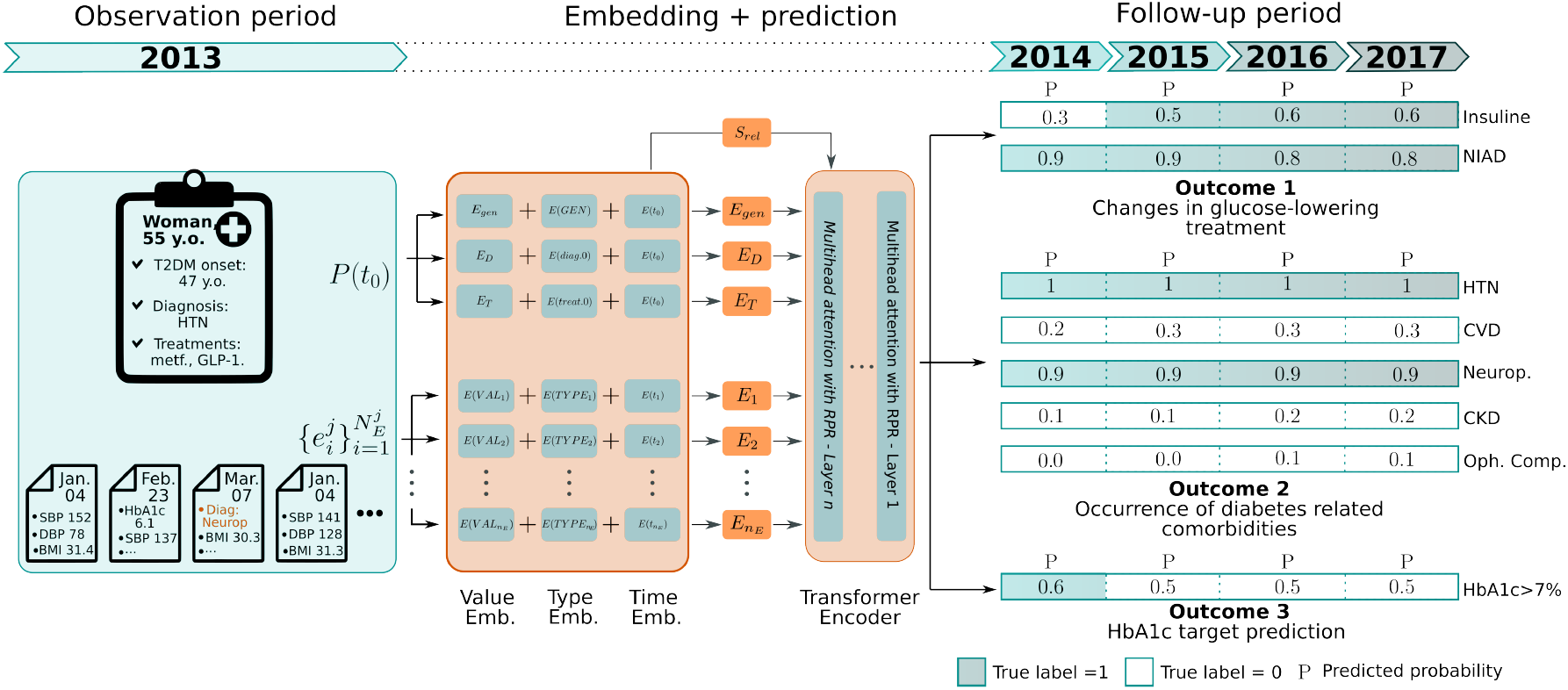
DARE basic architecture with prediction examples for the three outcomes. An embedding (emb) layer generates the *E*_*gen*_, *E*_*D*_, *E*_*T*_, and *E*_*i*_ vectors as the sum of the value embedding, type embedding and time embedding. These vectors feed the transformer encoder layer built with several attention layers with RPR. The vectors generated by the encoder are used for pre-training task, namely masked-language modeling (MLM) and Initial Status Prediction (ISP) tasks, and fine-tuning tasks. In this example DARE correctly predicts that the subject is prescribed with an insulin-based treatment during the second year of follow-up. At the same time, it correctly predicted no changes in the diagnosed comorbidities besides the diagnosis of neuropathy during the observation period. For the predicted HbA1c targets, the model correctly predicts the target of the first follow-up year, while the probability reduces for the following 3 years. (Abbreviations: y.o.=years old, HTN=Hypertension, metf.=Metformin, SBP/DBP=Systolic/diastolic blood pressure, BMI=Body mass index, NIAD=non-insulin diabetic drugs, Neurop.=Neuropathy, CVD=Cardiovascular diseases, Oph. Comp.=Ophthalmological complications, CKD=Chronic kidney disease)

This approach to input data provides two primary advantages: first, it enables the model to effectively process both dynamic data, such as sequences of measured variables, and static data, without requiring data imputation to fit them into a tabular format. Second, the use of an initial status vector *P*_*j*_(*t*_0_) restricts sequence length, which would otherwise grow excessively as time progresses. For clarity, we will omit the patient index *j* in further notation.

### 3.3. Model Pre-Training

In NLP, pre-trained large language models provide a key advantage: their adaptability to a range of tasks. This reduces both the time and data requirements for fine-tuning models on specific tasks, often leading to improving in performance. Common unsupervised pre-training tasks include Masked Language Modeling (MLM) and Next Sentence Prediction[26]. While MLM has been successfully adapted for use with other models on EHR data, NSP is less applicable to these data modalities. Therefore, to pre-train DARE, we used the traditional MLM task and introduced a novel pre-training task, Initial Status Prediction (ISP), aimed at predicting a patient’s initial diagnosis and medication usage.

For the MLM task, we proceeded as follows: we selected different measures with a probability *p* = 0.15, then we masked their values with a mask token in 70% of the cases, added white noise perturbation (*ϵ* ∼ *N*(0, 0.1)) in 15% of the cases, and left them as they were in the remaining cases. For each sequence of EHRs, the model has been trained to predict the values of the selected events.

For the ISP task, we masked the *P*_*D*_(*t*_0_) vectors with a probability *p* = 0.2, the *P*_*M*_(*t*_0_) vectors with a probability *p* = 0.2, and both vectors with a probability of *p* = 0.1. Ultimately, the model has been trained to predict the masked vectors.

We employed the Asynchronous Successive Halving Algorithm (ASHA)[27] to tune the hyper-parameters of the model, aiming to minimize the combined loss of the two pre-training tasks.

### 3.4. Outcomes Definition and Model Fine-tuning

The goal of the self-supervised pre-training was to produce contextualized embedding vectors for each data element in the sequence, allowing the model to capture the structural and temporal dependencies inherent in EHR data without directly predicting a specific outcome. This pretrained model can then be adapted to a variety of predictive tasks by adding a task-specific head and fine-tuning with limited labeled data, as the model has already learned general EHR patterns and relationships. Practically, to evaluate the utility of DARE and its adaptability to various clinically relevant tasks, we have chosen three prediction outcomes to be evaluated during the follow-up period (from the inclusion date to end of study in 2017), namely:

- Occurrence of diabetes-related comorbidities; we aimed at predicting new comorbidities within the subsequent 4 years.
- Changes in glucose-lowering treatment; our goal was to predict changes in the treatment. Specifically, we were interested in determining whether the subjects started using non-insulin diabetic drugs (NIAD, defined as presence of ATC/DDD codes: A10B and subgroups) or insulin (defined as presence of ATC/DDD codes: A10A and subgroups) in the following 4 years.
- Glycated hemoglobin (HbA1c) target prediction; we intended to predict whether subjects achieved target HbA1c levels in each of the following 4 years. A subject was considered to be within target during a year if their mean HbA1c level during that year was lower than 7%.[28]

For the three outcomes, we used a GRU based model to predict the longitudinal outputs from the embeddings generated by DARE.

### 3.5. Statistical analysis and Model Validation

Patients’ characteristics were reported using mean and SD for the clinical and analytical variables, and percentages for categorical variables. Fine-tuned models were trained using binary cross entropy loss and evaluated using the Area Under the receiver operating characteristic Curve (AUC)evaluated at each year of follow-up. Model comparisons were performed with 10-fold cross-validation. We employed the Welch Two Sample t-test with Bonferroni correction (*α* = 0.05) to statistically pairwise compare the performance of DARE against the three baseline models: a bidirectional GRU recurrent neural network, a random forest (RF) model and a logistic regression (LR). Moreover, to further validate the results, we fitted a linear model trying to study the differences between the GRU and DARE performances. For each one of the outcomes the model was:

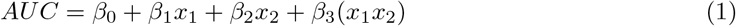

where *x*_1_ is the size of the training set and *x*_2_ indicates if the results were obtained with the GRU model.

All models were trained with PyTorch V1.12.1 [29] on two NVIDIA GeForce RTX 2080s. Statistical analysis used R (V3.6.3).

## 4. Results

Data of 232,885 people with T2DM from Catalunya (Spain) were extracted from the SIDIAP database. After applying the selection criteria described in Material and Methods section, we obtained 201,922 patients, 176,922 used for the pre-training phase and 25,000 for fine-tuning. Data preparation for the study is summarized in the graph in Supplementary Material, Figure S1, while baseline characteristics of study participants are reported in Supplementary Material, Table S2. Evolution of the outcomes during the follow-up period are shown in Table 1 while examples of the predicted outcomes together with the input sequence can be found in Figure 1 and in Supplementary Material, Figure S2.

**Table 1:** Outcomes events during the four years of follow-up period. Total number of events and percentages are reported.

|  | 2014 | 2015 | 2016 | 2017 |
| --- | --- | --- | --- | --- |
| **New Comorbidities** |  |  |  |  |
| Hypertension | 19338(77.35) | 20189(80.76) | 20793(83.17) | 20920(83.68) |
| Cardiovascular disease | 4022(16.09) | 4558(18.23) | 5093(20.37) | 5170(20.68) |
| Retinopathy | 3741(14.96) | 4189(16.76) | 4561(18.24) | 4830(19.32) |
| Chronic kidney disease | 1096(4.38) | 1289(5.16) | 1467(5.87) | 1526(6.10) |
| **New Antidiabetic treatments** |  |  |  |  |
| Insulin | 9533(38.13) | 10641(42.56) | 11510(46.04) | 11746(46.98) |
| Non-insulin | 22059(88.24) | 22012(88.05) | 21915(87.66) | 21570(86.28) |
| **Glycated hemoglobin (HbA1c)** |  |  |  |  |
| HbA1c,(%) mean (SD) | 7.55(1.32) | 7.57(1.31) | 7.56(1.31) | 7.51(1.27) |
| HbA1c < 7% | 10435(41.74) | 10168(40.67) | 10177(40.71) | 10404(41.62) |

### 4.1. Pre-training results

The hyper-parameter search yielded an optimal model configuration with three layers, eighteen attention heads, and a hidden size of 360. Details of the hyperparameters space explored may be found in the Supplementary Material, Table S3.

Evaluating embedding quality remains a challenging due to the lack of a universally accepted metric [30]. In Figure 2(a)-(b), we projected the vector representation of some clinical events from the training dataset onto a 2D space. Figure 2(a) shows a distinct stratification of the vectors into two separate areas which effectively distinguish patients with retinopathy from those without. Additionally, we observe clusters among patients with other comorbidities; for instance, patients with neuropathy tend to group together in the lower part of the plot while CVD are grouped in the upper part.

**Figure 2:**
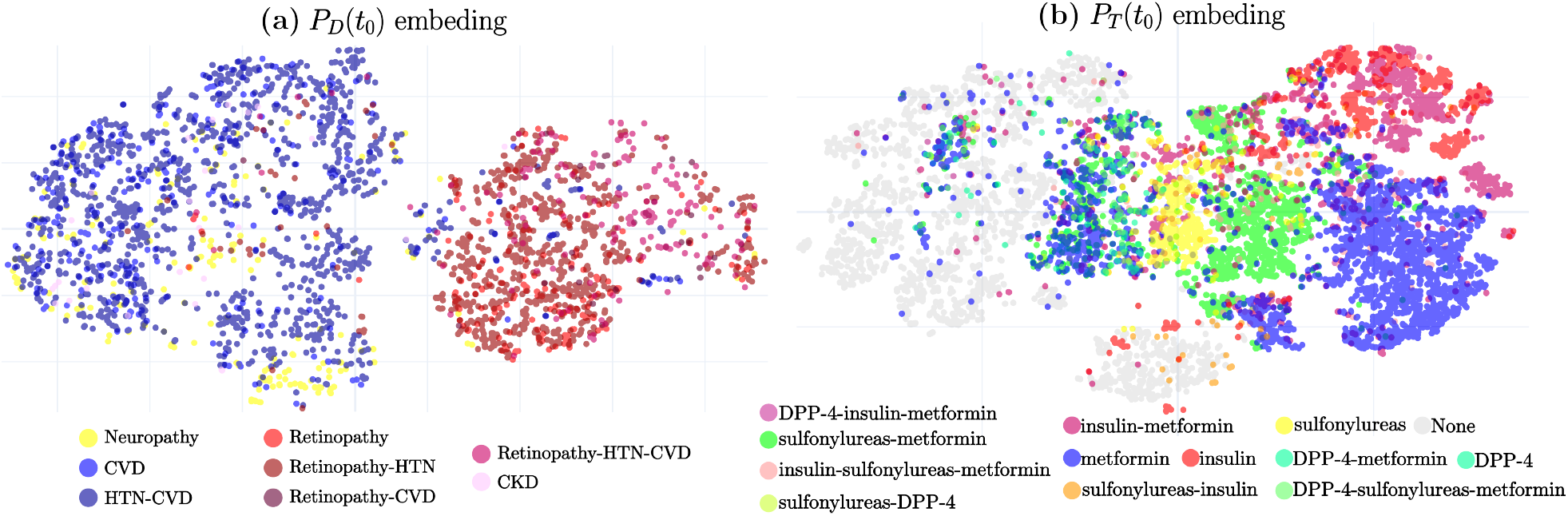
Visual inspection of the output space of DARE: each point represents embeddings of vectors *P*_*D*_(*t*_0_), and *P*_*T*_(*t*_0_) for 5000 random patients, with colors indicating their diagnoses and drug groups.

Figure 2(b) also reveals distinct clusters for medication embeddings. Patients without antidiabetic medication prescriptions tend to cluster in the lower left area, near embeddings for Metformin (the first-line treatment for T2DM [31]) and the combination of Metformin and Sulfonylureas (a common first-line add-on therapy [32]). Conversely, the top-right corner groups patients on insulin, typically used as a second- or third-line treatment for T2DM.

Additionally, in Supplementary Material, Figure S3, distinct trajectories of HbA1c evolution are depicted. HbA1c measurements from subjects who successfully maintained their HbA1c at a target of 7%, represented by square dots, are clustered in the upper and left portion of the plot. Conversely, measurements from patients who did not achieve HbA1c target levels exhibit greater dispersion. They move from the central region of the plot for measurements near the 7% target to the lower-right corner for higher HbA1c levels.

Another interesting property that differentiates attention-based models from other DL architectures is the possibility to investigate the learning patterns through a visual investigation of the self-attention weights. The attention patterns for three different heads in the first transformer layer are shown for three different subjects in Figure 3. In the sequence of data of the first patient, where attention connections for high creatinine value are highlighted, the different attention heads exhibit complementary behaviors: the green head focuses on variables measured close to the event (within a month), the blue head attends most strongly to variables measured before the event, and the orange head prioritizes connections with the initial status vectors. Interestingly, the blue head displays its strongest connections with other creatinine measurements and with measurements of the eGFR.

**Figure 3:**
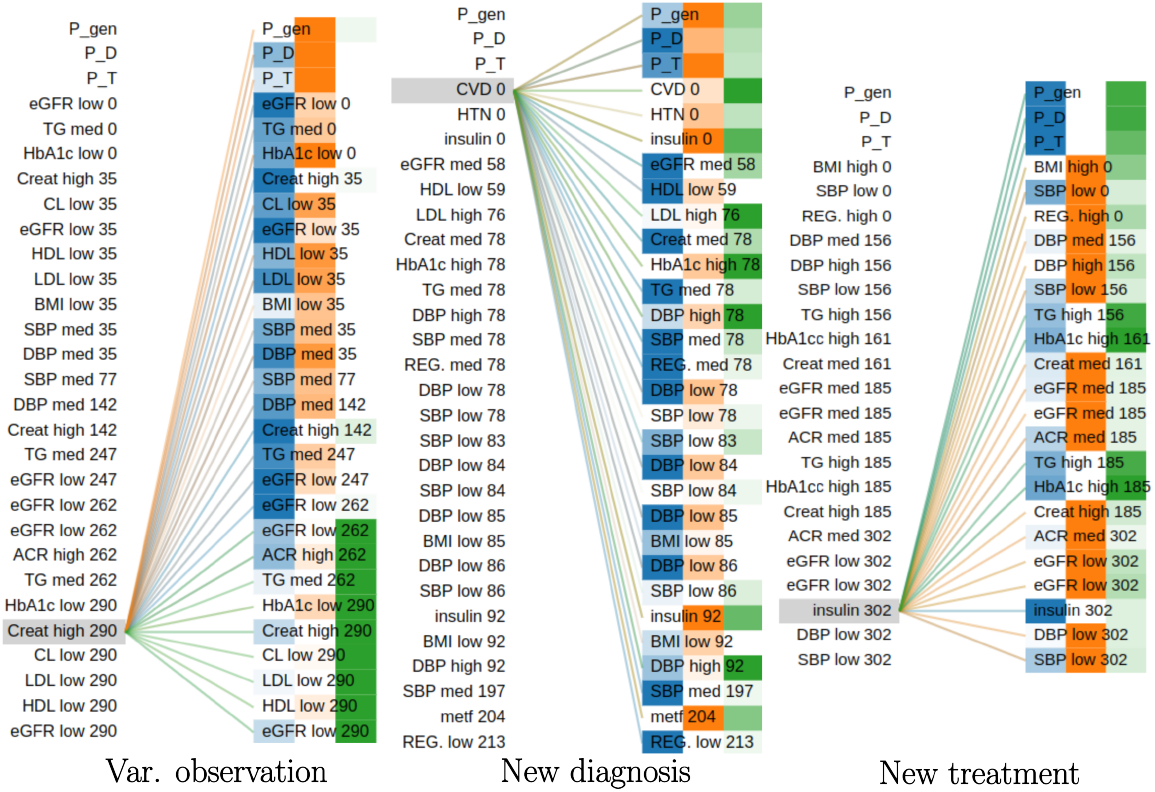
(Attention connections for different inputs and attention heads: colors indicate different heads while color intensity are proportional to attention weights.For high creatinine levels, while the green head focuses on recent measurements, the blue one dispalys the strongest connections with other creatinine and eGFR measures. In the cardiovascular example, heads emphasize event values, with the green head attending to high blood pressure and cholesterol values. For insulin prescriptions, attention highlights key markers like HbA1c and triglycerides, both critical for diabetes management. (Time from *t*_0_(in days) are indicated. For variables we included a label indicating if the measures was low(*v*_*var*_ *< µ −* 0.5*σ*), medium(med)(*µ −* 0.5*σ ≤ v*_*var*_ *≤ µ* + 0.5*σ*) or high(*v*_*var*_ *> µ* + 0.5*σ*))

In the second sequence, which illustrates attention connections related to a cardiovascular disease diagnosis, the heads appear to be more influenced by the values of the events rather than the timing of their occurrence. For example, the green head is predominantly activated by high values of blood pressure and cholesterol, both of which are typically associated with cardiovascular complications. In contrast, the blue head shows its strongest connections when these variables have medium or low values. Lastly, the orange head exhibits strong connections with the status vectors and the anti diabetic treatments.

In the final example, which illustrates attention connections for a new insulin prescription, the blue head demonstrates the strongest connections with the initial status vectors. Interestingly, the orange and green heads show a distinct pattern. The orange head prioritizes most measured variables, except for two. These two exceptions strongly activate the third head. Notably, the first exception is high HbA1c, a marker of poor diabetes control [33]. The second is high triglycerides, recently suggested as a target for diabetes management [33] and linked to insulin resistance [34].

### 4.2. Fine-tuning results

Fine-tuning results for the DARE model and the baseline models are reported in Table 2 and 3 for results stratified by sex and patients’ age for the two top performing models (DARE and GRU models). The estimated coefficients of the explanatory models are reported in table 4.

**Table 2:** DARE performances in validation tasks with 10-folds cross validation. Mean ROC-AUC(std). For each task, the ROC-AUC for each follow-up year is shown, along with the mean ROC-AUC for each predicted comorbidity/drugs class and the overall mean for the entire task.(*** for *p* < 0.001, * for 0.01 < *p* < 0.05,. for ≤ *p* < 0.1, Welch Two Sample t-test).

| (a) Comorbidities prediction task |  |  |  |  | (b) Antidiabetic treatments prediction task |  |  |  |  |
| --- | --- | --- | --- | --- | --- | --- | --- | --- | --- |
|  | DARE | GRU | RF | LR |  | DARE | GRU | RF | LR |
| Total | **0.88(0.00)\*\*\*** | 0.86(0.01) | 0.63(0.01) | 0.86(0.01) | Total | **0.91(0.00)\*\*\*** | 0.90(0.00) | 0.76(0.00) | 0.79(0.00) |
| HTN | **0.96(0.00)\*\*\*** | 0.94(0.01) | 0.76(0.01) | 0.94(0.00) | Insuline | **0.95(0.00)\*** | 0.94(0.00) | 0.87(0.00) | 0.87(0.01) |
| 2014 | **0.97(0.00)\*\*\*** | 0.95(0.01) | 0.93(0.00) | 0.96(0.00) | 2014 | **0.97(0.00)\*** | 0.96(0.00) | 0.91(0.00) | 0.91(0.00) |
| 2015 | **0.95(0.00)\*\*\*** | 0.94(0.01) | 0.80(0.00) | 0.94(0.00) | 2015 | **0.95(0.00)\*** | 0.94(0.00) | 0.87(0.00) | 0.87(0.00) |
| 2016 | **0.94(0.00)\*\*\*** | 0.92(0.01) | 0.66(0.00) | 0.92(0.00) | 2016 | **0.94(0.00)·** | 0.93(0.01) | 0.86(0.01) | 0.85(0.01) |
| 2017 | **0.94(0.01)\*\*\*** | 0.92(0.01) | 0.62(0.00) | 0.92(0.00) | 2017 | **0.92(0.00)\*** | 0.92(0.01) | 0.84(0.01) | 0.84(0.01) |
| CVD | **0.87(0.01)·** | 0.86(0.02) | 0.57(0.01) | 0.85(0.01) | NIAD | **0.88(0.01)\*** | 0.87(0.01) | 0.64(0.01) | 0.70(0.01) |
| 2014 | **0.92(0.01)\*** | 0.90(0.01) | 0.57(0.00) | 0.91(0.01) | 2014 | **0.92(0.01)\*** | 0.91(0.01) | 0.74(0.01) | 0.83(0.01) |
| 2015 | **0.88(0.01)\*** | 0.86(0.02) | 0.57(0.00) | 0.87(0.01) | 2015 | **0.89(0.01)\*** | 0.87(0.01) | 0.66(0.01) | 0.71(0.01) |
| 2016 | **0.84(0.02)** | 0.83(0.02) | 0.57(0.00) | 0.82(0.01) | 2016 | **0.86(0.01)·** | 0.85(0.01) | 0.61(0.01) | 0.65(0.01) |
| 2017 | **0.83(0.02)** | 0.82(0.02) | 0.57(0.00) | 0.80(0.01) | 2017 | **0.84(0.01)\*** | 0.82(0.01) | 0.58(0.01) | 0.62(0.01) |
| Neuropathy | **0.79(0.01)\*** | 0.78(0.01) | 0.54(0.01) | 0.77(0.01) |  |  |  |  |  |
| 2014 | 0.84(0.01) | 0.81(0.01) | 0.53(0.00) | **0.86(0.01)\*** |  |  |  |  |  |
| 2015 | 0.78(0.01) | 0.77(0.01) | 0.55(0.00) | **0.79(0.02)** |  |  |  |  |  |
| 2016 | **0.74(0.01)** | **0.74(0.01)** | 0.56(0.00) | **0.74(0.01)** |  |  |  |  |  |
| 2017 | **0.73(0.01)** | **0.73(0.01)** | 0.55(0.00) | 0.72(0.01) |  |  |  |  |  |
| Retinopathy | **0.91(0.00)\*\*\*** | 0.89(0.01) | 0.71(0.02) | 0.86(0.01) |  |  |  |  |  |
| 2014 | **0.95(0.01)\*\*\*** | 0.92(0.01) | 0.70(0.00) | 0.92(0.01) |  |  |  |  |  |
| 2015 | **0.92(0.01)\*\*\*** | 0.89(0.01) | 0.71(0.00) | 0.88(0.00) |  |  |  |  |  |
| 2016 | **0.89(0.01)\*\*\*** | 0.87(0.01) | 0.70(0.00) | 0.84(0.01) |  |  |  |  |  |
| 2017 | **0.88(0.01)\*\*\*** | 0.86(0.01) | 0.71(0.00) | 0.82(0.01) |  |  |  |  |  |
| CKD | **0.88(0.01)\*** | 0.87(0.01) | 0.51(0.00) | 0.85(0.01) |  |  |  |  |  |
| 2014 | **0.93(0.01)\*\*\*** | 0.91(0.01) | 0.50(0.00) | 0.90(0.01) |  |  |  |  |  |
| 2015 | **0.89(0.01)\*** | 0.87(0.01) | 0.50(0.00) | 0.86(0.01) |  |  |  |  |  |
| 2016 | **0.86(0.01)** | 0.85(0.01) | 0.51(0.00) | 0.83(0.01) |  |  |  |  |  |
| 2017 | **0.85(0.02)** | 0.84(0.01) | 0.51(0.00) | 0.82(0.01) |  |  |  |  |  |

| (c) HbA1c target prediction task |  |  |  |  |
| --- | --- | --- | --- | --- |
|  | DARE | GRU | RF | LR |
| Total | **0.82(0.01)** | **0.82(0.01)** | 0.73(0.01) | 0.73(0.01) |
| 2014 | **0.88(0.01)** | **0.88(0.01)** | 0.79(0.01) | 0.79(0.02) |
| 2015 | **0.83(0.01)** | **0.83(0.01)** | 0.74(0.01) | 0.74(0.01) |
| 2016 | **0.80(0.01)** | **0.80(0.01)** | 0.71(0.01) | 0.71(0.01) |
| 2017 | 0.77(0.01) | **0.78(0.01)** | 0.69(0.01) | 0.69(0.01) |

**Table 3:** Performances of the top two models stratified by sex and age. Age stratified by quartiles, mean ROC-AUC(std) (*** for *p* < 0.001, * for 0.01 < *p* < 0.05, · for 0.5 ≤ *p* < 0.1, Welch Two Sample t-test).

| (a) DARE |  |  |  |  |  |  |  |  |
| --- | --- | --- | --- | --- | --- | --- | --- | --- |
|  | Age<64 years |  | Age<73 years |  | Age<80 years |  | Age>80 years |  |
|  | F<br>N=2828 | M<br>N=4492 | F<br>N=3582 | M<br>N=2914 | F<br>N=1989 | M<br>N=1246 | F<br>N=3609 | M<br>N=4340 |
| HTN | **0.95(0.01)\*** | **0.94(0.01)\*\*\*** | **0.97(0.01)\*** | **0.96(0.01)\*** | **0.97(0.02)** | **0.96(0.02)** | **0.96(0.01)\*** | **0.95(0.01)\*** |
| CVD | **0.84(0.04)** | **0.87(0.02)\*** | **0.85(0.03)** | **0.89(0.02)** | **0.84(0.04)** | **0.89(0.03)** | **0.84(0.04)** | **0.87(0.02)** |
| Neuropathy | **0.81(0.02)\*** | **0.78(0.02)** | **0.77(0.03)** | **0.75(0.03)** | **0.79(0.04)\*** | **0.77(0.06)** | **0.80(0.02)** | **0.79(0.03)** |
| Retinopathy | **0.91(0.02)\*\*\*** | **0.91(0.01)\*\*\*** | **0.91(0.02)·** | **0.91(0.02)** | **0.93(0.02)** | **0.92(0.04)** | **0.92(0.02)·** | **0.91(0.02)\*** |
| CKD | **0.89(0.07)** | **0.88(0.03)** | **0.88(0.04)** | **0.88(0.05)** | **0.86(0.08)** | **0.82(0.08)** | **0.90(0.04)** | **0.88(0.03)** |
| Insuline | **0.94(0.01)** | **0.94(0.01)** | **0.95(0.00)** | **0.95(0.01)** | **0.94(0.01)·** | **0.94(0.01)·** | **0.96(0.01)** | **0.95(0.01)** |
| NIAD | **0.85(0.03)** | **0.86(0.05)** | **0.87(0.02)·** | **0.88(0.03)** | **0.85(0.03)** | **0.85(0.02)** | **0.88(0.03)** | **0.89(0.02)·** |
| HbA1c target | **0.83(0.02)** | **0.81(0.01)** | **0.83(0.01)** | **0.82(0.02)** | **0.82(0.01)** | **0.81(0.03)** | **0.84(0.01)** | **0.82(0.01)** |

| (b) GRU |  |  |  |  |  |  |  |  |
| --- | --- | --- | --- | --- | --- | --- | --- | --- |
|  | Age<64 years |  | Age<73 years |  | Age<80 years |  | Age>80 years |  |
|  | F<br>N=2828 | M<br>N=4492 | F<br>N=3582 | M<br>N=2914 | F<br>N=1989 | M<br>N=1246 | F<br>N=3609 | M<br>N=4340 |
| HTN | 0.93(0.01) | 0.91(0.01) | 0.96(0.01) | 0.94(0.02) | 0.96(0.02) | 0.94(0.02) | 0.95(0.01) | 0.93(0.01) |
| CVD | 0.82(0.05) | 0.85(0.02) | 0.84(0.02) | 0.88(0.03) | 0.82(0.06) | 0.87(0.03) | 0.83(0.04) | 0.86(0.02) |
| Neuropathy | 0.78(0.02) | 0.77(0.03) | **0.77(0.02)** | **0.75(0.03)** | 0.74(0.03) | 0.75(0.06) | 0.79(0.02) | 0.78(0.02) |
| Retinopathy | 0.87(0.02) | 0.87(0.02) | 0.89(0.03) | 0.90(0.03) | 0.91(0.02) | 0.90(0.05) | 0.90(0.03) | 0.88(0.03) |
| CKD | 0.87(0.08) | 0.87(0.03) | 0.86(0.07) | 0.87(0.04) | 0.81(0.12) | **0.82(0.08)** | 0.88(0.03) | 0.86(0.03) |
| Insuline | **0.94(0.01)** | 0.93(0.01) | 0.95(0.01) | 0.94(0.01) | 0.93(0.01) | 0.93(0.01) | 0.95(0.01) | 0.95(0.01) |
| NIAD | 0.83(0.04) | 0.84(0.04) | 0.86(0.02) | 0.87(0.02) | 0.84(0.03) | **0.85(0.03)** | 0.86(0.02) | 0.88(0.02) |
| HbA1c target | **0.83(0.02)** | **0.81(0.01)** | **0.83(0.01)** | **0.82(0.02)** | **0.82(0.02)** | **0.81(0.02)** | **0.84(0.01)** | **0.82(0.01)** |

**Table 4:** Estimated regression coefficients for the explanatory models of the three tasks. A negative value for the *β*_2_ coefficient indicates an improvement with respect to the baseline model (GRU). A positive *β*_3_ coefficient indicates that the baseline model is more sensible to the training set size.

| | $\beta_0$<br>intersect | $\beta_1$<br>N. pats. | $\beta_2$<br>GRU | $\beta_3$<br>GRU.N.pats. |
| --- | --- | --- | --- | --- |
| Comorbidities | 8.0e-1*** | 4.0e-6*** | 9.8e-3* | -1.4e-6*** |
| Treatments | 8.9e-1*** | 1.1e-6*** | -2.8e-2*** | 9.5e-7*** |
| Targets | 8.1e-1*** | 7.8e-7 | -8.9e-2*** | 4.7e-6*** |

N. pats.: Number of Patients in train
\*\*\* for $p < 0.001$ , \* for $0.01 < p < 0.05$ , · for $0.5 \leq p < 0.1$

DARE’s embeddings consistently improved the performance of the prediction layers across all three tasks. Notably, for predicting diabetes-related comorbidities and glucose-lowering treatments, DARE achieved statistically significant improvement over the GRU model (second-best) in the 10-fold cross-validation. This positive effect held true for most diagnosis and prediction years.

For predicting glucose-lowering treatments (shown in Supplementary Material, Figure S4(a)), DARE’s performance was statistically better than other models regardless of training set size. Interestingly, the performance gap between DARE and the GRU model (second-best) widened as the training data size decreased. This suggests that the GRU model is more sensitive to limited training data, as confirmed by the estimated regression coefficients *β*_2_ and *β*_3_. These coefficients indicate that the GRU model has a lower overall AUC and is more susceptible to reductions in AUC when the training data size shrinks.

DARE’s performance on predicting diabetes-related comorbidities showed a different trend (Supplementary Material, Figure S4(b)). Its performance degraded faster for smaller training sets, particularly when fewer than 10,000 patients were used for training. In contrast, the performance for predicting HbA1c target levels (shown in Supplementary Material, Figure S4(c)) remained relatively stable even with smaller training sets. The estimated regression coefficient *β*_1_ was not statistically significant, indicating minimal influence of training set size on predictions for this task. Conversely, the GRU model’s performance suffered significantly when fewer patients were available for training, as confirmed by the estimated *β*_3_ coefficient.

## 5. Discussion and Conclusions

In this study, we introduced an encoder model based on the Transformer encoder architecture(DARE) that leverages the transfer learning paradigm to enhance performances on various clinically relevant outcomes in individuals with T2DM.

DARE’s architecture is designed to handle different data types. It can represent diagnosis codes, medication prescriptions, and clinical/analytical measurements within a unified embedding space. This approach is highly flexible and potentially applicable to a broad range of clinical data. Each record in the EHR sequence is initially embedded as a triplet considering time, type, and value of the record. This versatile concept can be applied to various clinical data formats. In addition to the EHR sequence, DARE incorporates static data like sex, age at diagnosis, and a summary of the patient’s health status at the beginning of the sequence. This strategy allows the model to limit the sequence length, focusing on recent events while retaining information about the patient’s medical history.

Visual analysis of the embedding vectors suggests that during the pre-training phase, the model successfully learned the underlying structure of the ICD-10 and ATC-DDD ontologies. DARE effectively mapped related codes together, grouping ICD-10 codes associated with the same comorbidities and ATC-DDD codes representing medications within the same pharmacological class. Furthermore, the investigation of the attention patterns learned during the pre-training phase shows that the model learned the relationship between different codes and variables measures. Similar behaviors where observed also in other pre-trained models for EHRs, with attention patterns showing strong connection between diseases and corresponding medications or even future comorbidity [14, 13].

When applied to predicting changes in glucose-lowering treatments, DARE significantly outperformed baseline methods. Notably, DARE delivered more consistent results compared to other deep learning approaches, regardless of the number of patients included in the fine-tuning training set. This suggests that DARE is less susceptible to variations in training data size.

The same behaviour was found for the prediction of HbA1c targets within four years after the last event. In this scenario, the impact of the training set size on HbA1c target prediction appears to be minimal (p*>*0.1). For the prediction of the most common diabetes comorbidities, the DARE model struggled to maintain a stable performance as the training set size decreased. We hypothesize that this is due to the imbalanced nature of the data: there are significantly more patients without new diagnoses compared to those who receive new ones. As the training set shrinks, DARE struggles to learn effectively from the sequence of EHR events. In contrast, the GRU model appears to simply copy the diagnosis information from the initial patient status vector *P*_*D*_(*t*_0_) instead of learning from the sequence. To investigate this hypothesis, we trained both models on an extremely small dataset (2,500 patients) with the EHR sequences masked and only the initial status data *P*(*t*_0_) included. The results suggrests to support our hypothesis: DARE’s AUC score decreased from 0.78(0.01) to 0.74(0.02). This indicates that DARE relies on the event sequence for accurate prediction. Conversely, the GRU model’s AUC score slightly increased from 0.80(0.01) to 0.81(0.01). This suggests that the GRU model primarily memorizes the initial diagnosis information (which might be sufficient for a small imbalanced dataset).

Despite the encouraging results shown in this work, there are various improvements that we aim to develop in the future and other properties to investigate. While the pre-training dataset included information for over 200,000 patients over 5 years, it remains limited compared to similar studies. In future work, we aim to retrain the DARE model with even larger datasets to investigate how the pre-training set size impacts fine-tuning performance. Even though this study focused on type 2 diabetes, the proposed approach has the potential to be applied to understanding the evolution of various chronic diseases. We plan to include data for different chronic diseases in the dataset to broaden the applicability of our approach.

In conclusion, this work introduces a novel deep learning encoder model based on the Transformer architecture that effectively analyzes Electronic Health Records (EHR) data. Unlike other DL models, DARE can learn the complex relationships between different data modalities. To do so, we introduced a new formalism for the representation of long sequences of clinical data, representing recent RCHCD as events and condensing previous information into initial status vectors *P*(*t*_0_). DARE demonstrates significant improvements in predicting different health outcomes, while also reducing the requirement for large, labeled training datasets - a major hurdle for many deep learning models. This characteristic allows DARE to be fine-tuned for diverse clinically relevant tasks, potentially paving the way for the development of novel preventive healthcare strategies.

## Data Availability

The data analysed in this study is subject to the following licenses/restrictions: restrictions apply to the availability of some or all data generated or analysed during this study because they were used under license. The corresponding authors will on request detail the restrictions and any conditions under which access to some data may be provided.

## Data availability

DARE code is open source and available at https://github.com/enriminzo/DARE

## Acknowledgments

This work was supported by the Grant PID2021-122952OB-I00 funded by AEI 10.13039/501100011033 and by ERDF A way of making Europe; the Networking Biomedical Research Centre in the subject area of Bioengineering, Biomaterials and Nanomedicine (CIBER-BBN), initiatives of Instituto de Investigación Carlos III (ISCIII); ISCIII (grant AC22/00035); and the CERCA Programme / Generalitat de Catalunya. B2SLab is certified as 2021 SGR 01052. This study was possible thanks to the commitment of physicians and nurses working in the Catalan Health Institute to provide optimal care to patients with diabetes. CIBER of Diabetes and Associated Metabolic Diseases (CIBERDEM) is an initiative from Instituto de Salud Carlos III, Madrid, Spain.

This analysis is part of the DiaCare Project of Novo Nordisk and the Fundació TicSalut (Departament de Salut, Generalitat de Catalunya), in collaboration with Evidenze Health España, for the benefit of people with type 2 diabetes.

## Appendix A. Model implementation details

### Input representation

The model input consists of an initial status vector *P*(*t*_0_) ideally composed of three vectors (*P*_*gen*_,*P*_*D*_(*t*_0_), *P*_*T*_(*t*_0_)) representing general static information of the patient, diagnosis, and treatments:

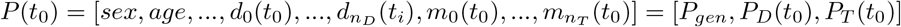

and a series of clinical of routinely collected health-care data 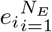 with *e*_*i*_ = (*TY PE*_*i*_, *V AL*_*i*_, *t*_*i*_) where:

- *TY PE*_*i*_ indicates if the health-care data is a diagnosis (*d*), a prescription of a pharmacological treatment (*t*), or an observation of one of the analytical or clinical variables (*var*).
- *V AL*_*i*_ is the value of the event, that can be represented with 3 vectors:
  — 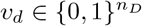, the 1-H encoding of a new diagnosis.
  — 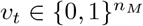, the 1-H encoding of a new glucose-lowering prescription.
  — 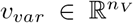, representing the measured variables. Note that to speed up training, numerical data have been standardized before being used as input of the model.
- *t*_*i*_ is the time of the event, expressed in days from *t*_0_.

Note that we used only sequences that had 5 or more data points in the same year, (one of these being a measure of HbA1c).

### Embedding layer

Inputs are mapped by an embedding layer in a series of vectors {*E*_*gen*_, *E*_*D*_, *E*_*T*_, *E*_1_, *E*_2_, …, *E*_*N*_*E*} with 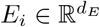, each one composed by the sum of three parts:

- **Value embedding**: for an event *e*_*i*_ the value embedding is *E*(*V AL*_*i*_) = *V*_*d*_ *× v*_*d*_ + *V*_*t*_ *× v*_*t*_ + *tanh*(*V*_*var*_ *×v*_*var*_) (note that just one of the tree vectors will be no zero vector) with *V*_*d*_, *V*_*t*_ and *V*_*var*_ parameters to learn within the model. We embedded the status vector divided by the 3 vectors that composed it, obtaining 3 embedding: *E*(*P*_*gen*_) = *V*_*gen*_ *×P*_*gen*_, *E*(*P*_*D*_) = *V*_*d*_ *×P*_*D*_, *E*(*P*_*T*_) = *V*_*t*_ *× P*_*T*_, with *V*_*gen*_ parameters to learn within the model, while *V*_*d*_ and *V*_*m*_ are the same for the status vectors and the events.
- **Type embedding**: obtained with a lookup table that map the TYPE token in a vector of the same dimension of the embedding space (*n*_*E*_).
- **Time embedding**: we used a fully-learnable time representation [35] as it has been show it may lead to better results in classification tasks compared to the classical fixed sinusoidal embedding of the BERT model [36].

### Attention Encoder

The original BERT model was composed by a stack of transformer layer.[11] The Core of the transformer is the multi-head attention mechanism: given an input sequence of vectors 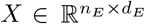, it transforms them into queries *Q* = *XW*_*Q*_, keys *K* = *XW*_*K*_ and values *V* = *XW*_*V*_ with *W*_*Q*_, *W*_*R*_, 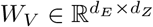 learnable parameters. Queries and keys vectors are then used to calculate the attention weights to be multiplied by the values vectors. Hence, the output is calculated as:

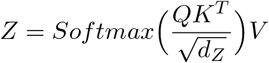

Attention with Relative Position Representations [9] introduced a new term in the calculation of the weights to take into account the distance between the corresponding vectors, namely:

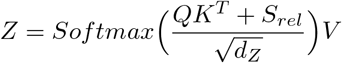

Where each element of the matrix 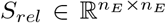 is calculated as *s*_*i,j*_ = *q*_*i*_(*a*_*i,j*_)^*T*^, being *q*_*i*_ the query corresponding to the i-th element in the input sequence and 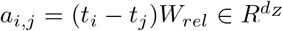 the embedding of the relative distance between events i and j.

## Appendix B. Supplementary figures

**Figure B.4:**
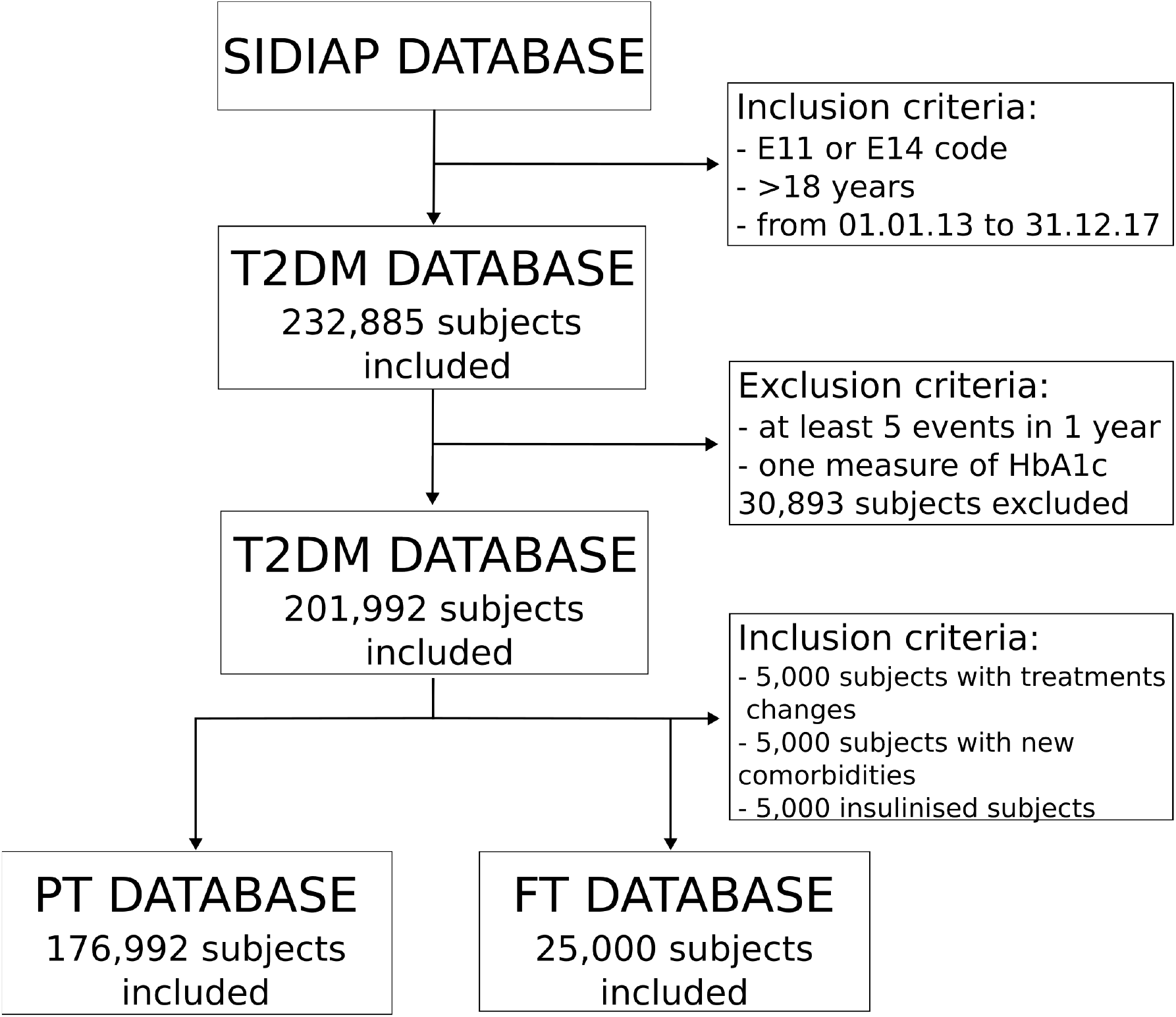
Study subject diagram. (Abbreviations: PT =Pre-training, FT=Fine-tuning).

**Figure B.5:**
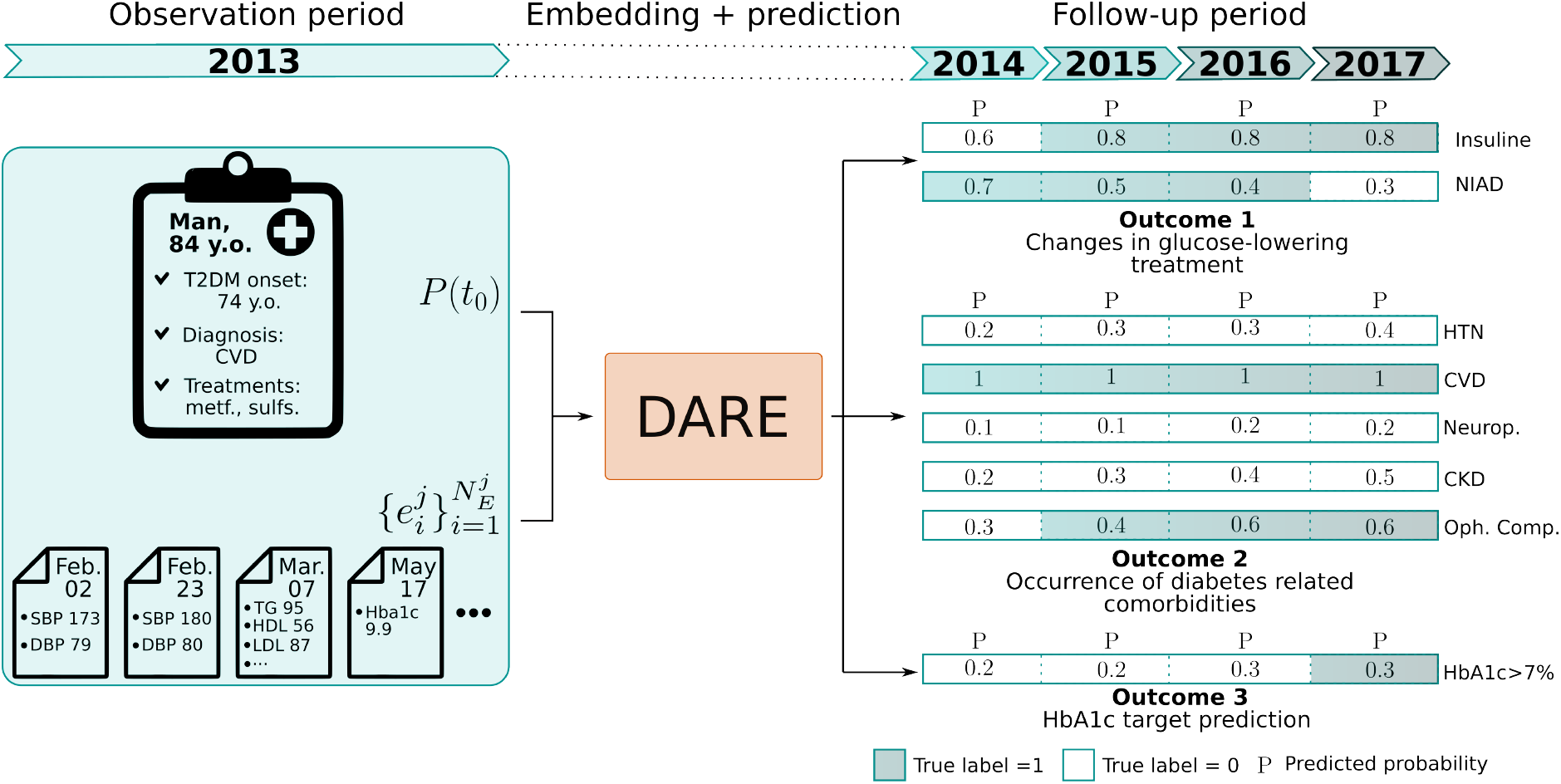
Fine tuning prediction example for the three outcomes. The model correctly predicts the change in treatment of the patient, moving from non-insulin to insulin base glucose-lowering treatment. Similarly, it predicts the CKD onset, even if it is a year late. Finally, even if it fails to correctly predict that the patient reaches HbA1c target in the 4th year of follow-up, the predicted target probability has a positive trend, showing that the model tends to predict an improvement of the HbA1c levels. (Abbreviations: y.o.=years old, CVD=Cardiovascular disease, metf.=Metformin, sulfs.= Sulfonylurea, SBP/DBP=Systolic/diastolic blood pressure, TG=triglycerides, HDL= High density lipoprotein, LDL=Low density lipoprotein, HbA1c=glycated haemoglobin, NIAD=non-insulin diabetic drugs, Neurop.=Neuropathy, HTN=Hypertension, Retinop.=Retinopathy, CKD=Chronic kidney disease)

**Figure B.6:**
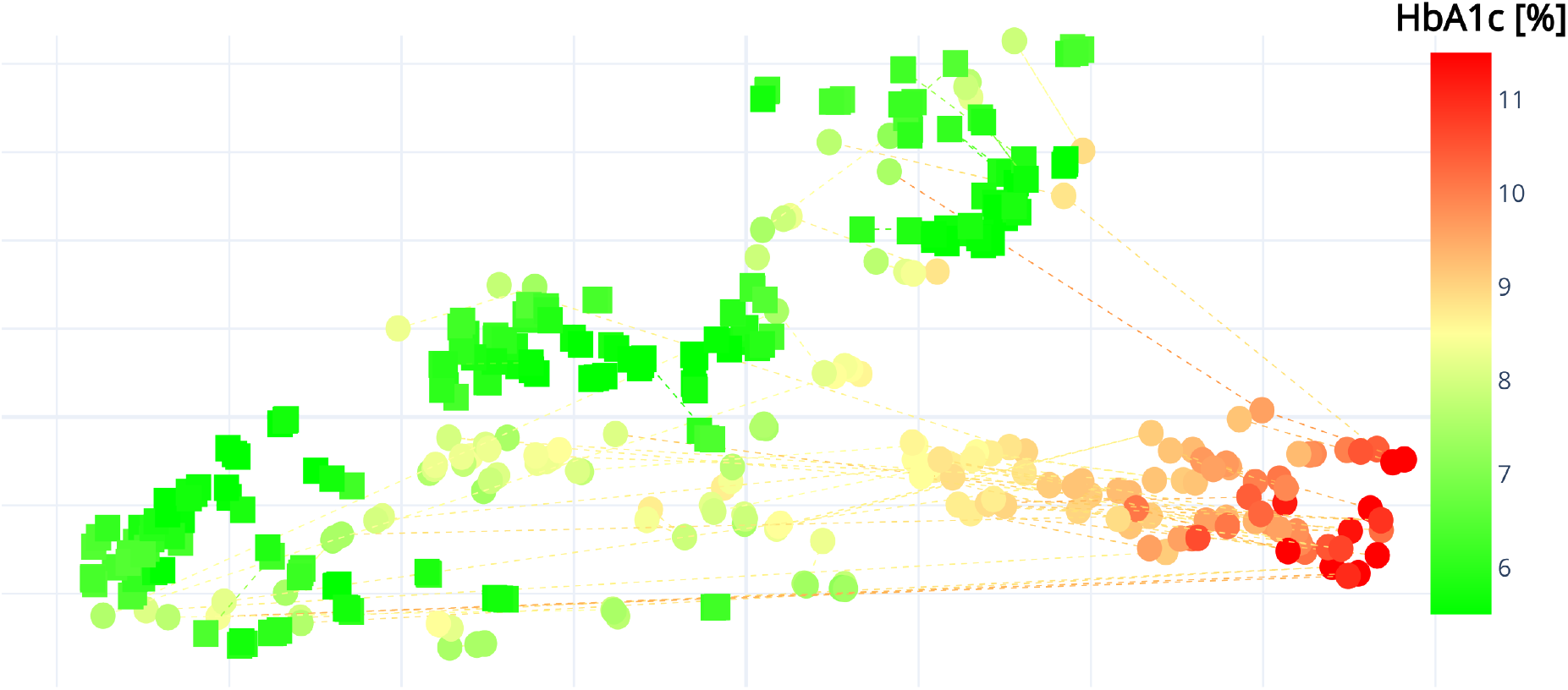
Visual inspection of the output space of DARE. Each point represents a different HbA1c measurement, with dotted lines connecting consecutive measurements of the same patient. Colours represent different HbA1c levels, while the shapes of the dots distinguish between subjects who consistently maintain their target HbA1c level (squares) and those who do not (circles).

**Figure B.7:**
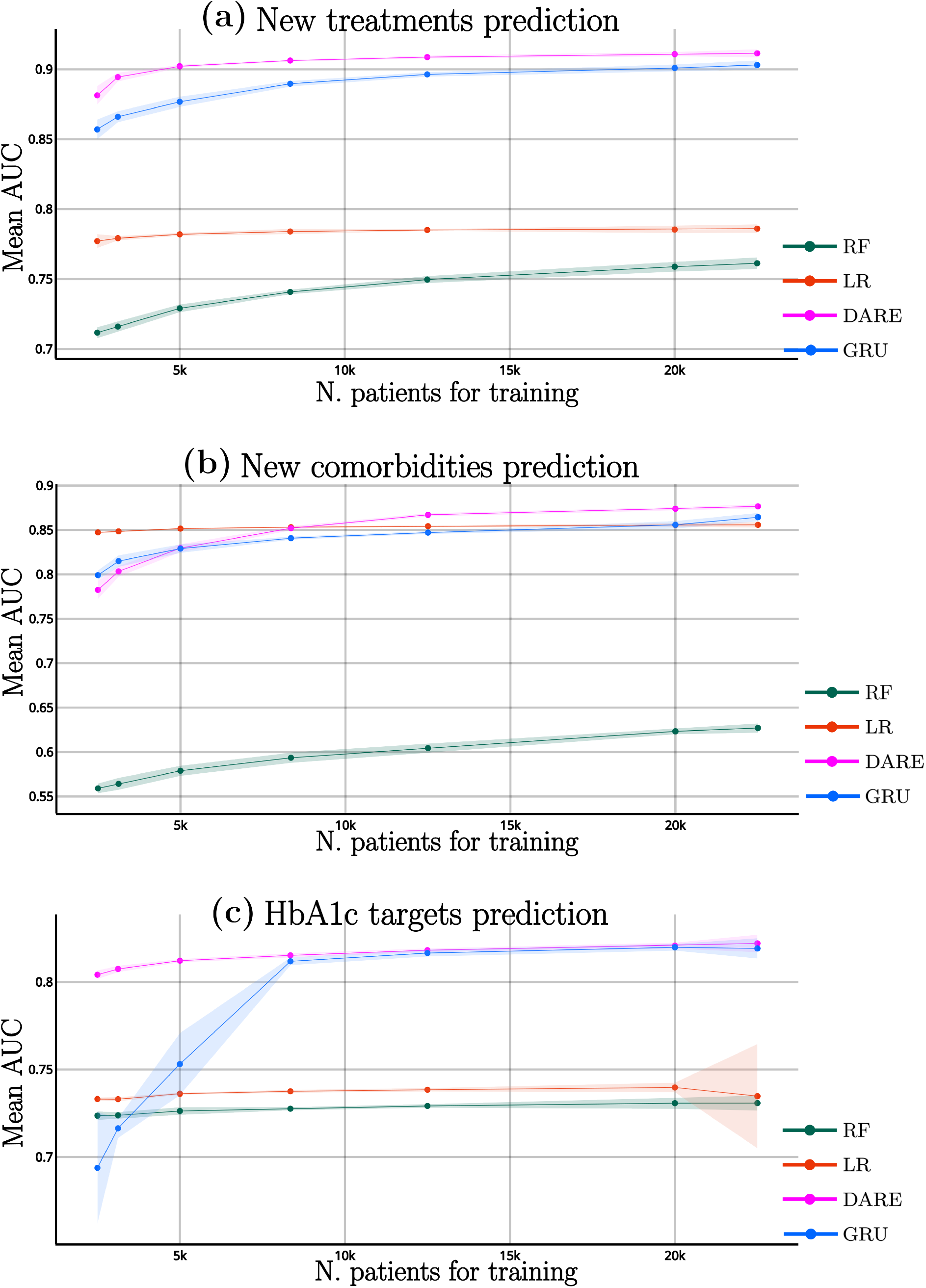
DARE results on fine-tuning. Mean AUC for the fine-tuning tasks for varied sizes of the training set. The lighter intervals indicate 95% confidence intervals.

## Appendix C. Supplementary tables

**Table C.5:** ICD-10 codes with the corresponding comorbidities groups.

| comorbidity | abbreviation | codes |
| --- | --- | --- |
| Hypertension | HTN | I10 I11 I11.0 I11.9 I12 I12.0 I12.9 I13 I13.0 I13.1 I13.2 I13.9 I15 I15.0 I15.1 I15.2 I15.8 I15.9 |
| Chronic vascular diseases | CVD | I20 I20.0 I20.17 I20.8 I20.9 I21 I21.0 I21.1 I21.2 I21.3 I21.4 I21.9 I22 I22.0 I22.1 I22.8 I22.9 I23 I23.0 I23.1 I23.2 I23.3 I23.4 I23.6 I23.8 I24 I24.0 I24.1 I24.8 I24.9 I25 I25.0 I25.1 I25.2 I25.3 I25.4 I25.5 I25.6 I25.8 I25.9 T82.2 E11.5 Z95.1 Z95.5 |
| Neuropathies and neurological complications |  | E11.4 E14.4 G13.0 G50 G50.0 G50.17 G50.8 G50.9 G51 G51.0 G51.1 G51.2 G51.3 G51.4 G51.8 G51.9 G52 G52.1 G52.2 G52.3 G52.7 G52.8 G52.9 G53 G53.0 G53.1 G53.3 G53.8 G54 G54.0 G54.1 G54.2 G54.3 G54.4 G54.5 G54.6 G54.7 G54.8 G54.9 G55 G55.0 G55.1 G55.2 G55.3 G55.8 G56 G56.0 G56.1 G56.2 G56.3 G56.4 G56.8 G56.9 G57 G57.0 G57.1 G57.2 G57.3 G57.4 G57.5 G57.6 G57.8 G57.9 G58 G58.0 G58.7 G58.8 G58.9 G59 G59.0 G59.8 G60 G60.0 G60.17 G60.17 G60.8 G60.9 G61 G61.0 G61.1 G61.8 G61.9 G62 G62.0 G62.1 G62.2 G62.8 G62.9 G63 G63.0 G63.2 G63.3 G63.5 G63.6 G63.8 G64 |
| Opthamological complications |  | E11.3 E13.3 E14.3 H36.0 |
| Chronic kidneys diseases | CKD | E11.2 N08.3 E13.2 E14.2 |

**Table C.6:**
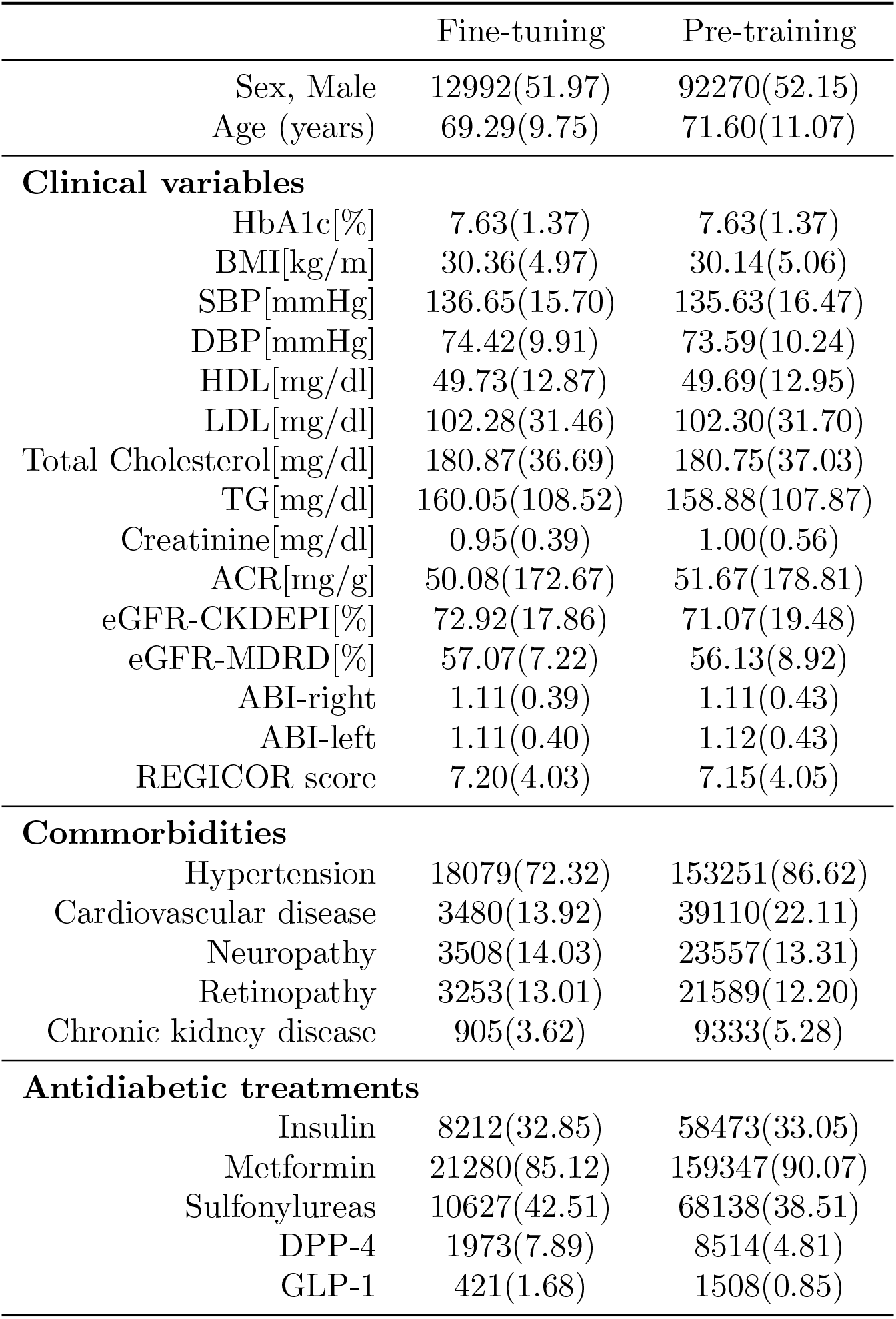
Basal values for pre-training and fine-tuning data. Data are reported as mean and standard deviation for continuous variables and with number of events and percentages for categorical ones. (Abbreviations: HbA1c=Glycated haemoglobin, BMI=Body mass index, SBP/DBP= Systolic/diastolic blood pressure, HDL=High density lipoprotein, LDL= Low density lipoprotein, TG=Triglycerides, ACR=Albumin/creatinine ratio, eGFR=estimated glomerular filtration rate, ABI=ankle brachial index, DPP-4=Dipeptidyl peptidase-4 inhibitor, GLP-1 = Glucagon-like peptide-1 receptor agonists)

**Table C.7:**
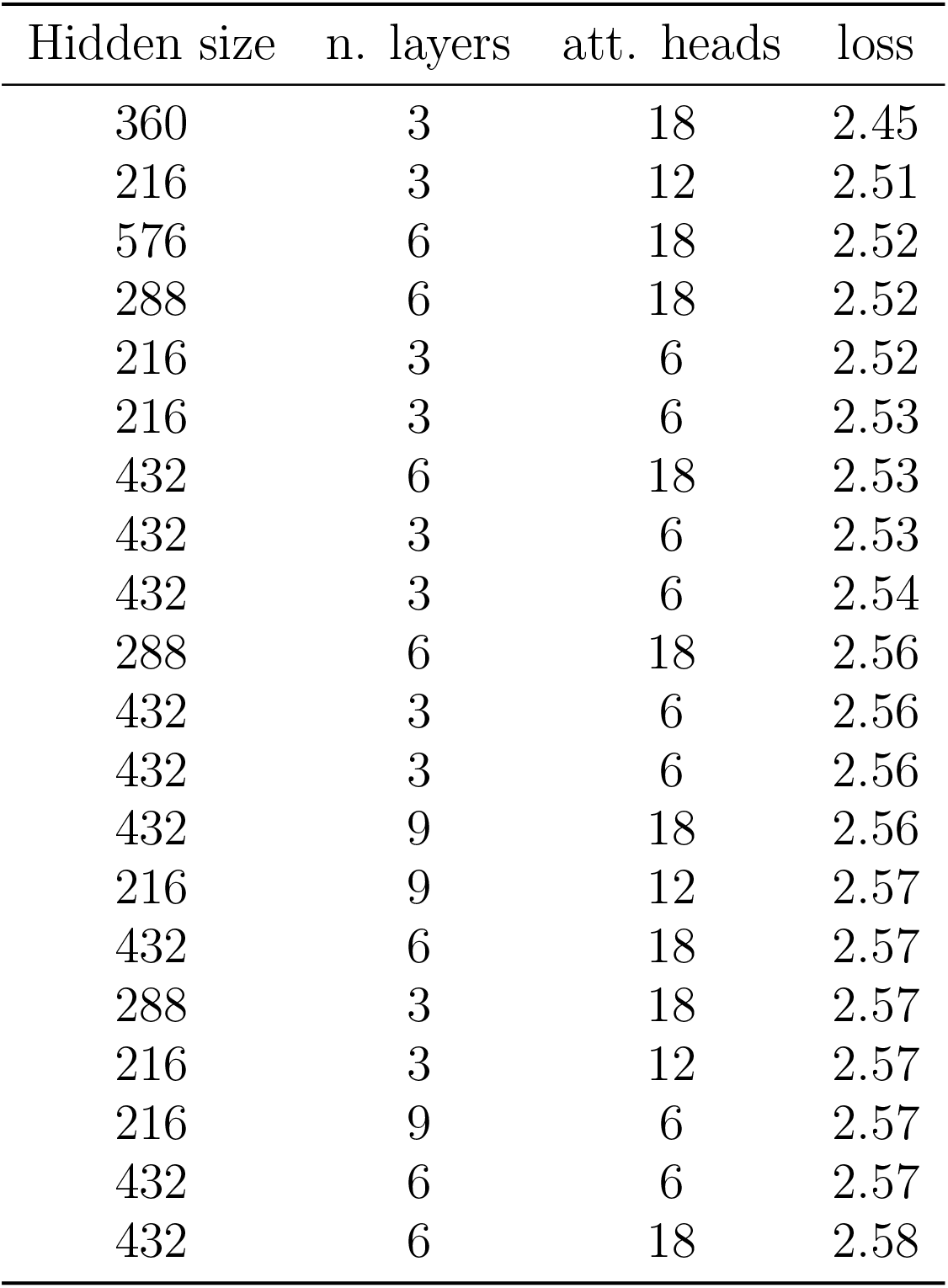
Results of the parameter search algorithm for the top 20 configurations. Explored hyperparemeters where: hidden size: [216, 288, 360, 432, 576]; number of layers: [3, 6, 9]; attention heads: [6, 12, 18].

